# How do the activities of daily living decline in people living with rarer dementias? A systematic review

**DOI:** 10.1101/2022.09.22.22280192

**Authors:** Beatrice Taylor, Suraya Mohamud, Emilie V Brotherhood, Emma Harding, Claire Waddington, Paul M Camic, Daniel C Alexander, Sebastian J Crutch, Joshua Stott, Chris Hardy, Neil P Oxtoby

## Abstract

**Objective:** To study how activities of daily living (ADLs) decline over the progressive course of rarer dementias (prevalence below 10%), in a systematic review of the literature.

Methods Relevant studies were identified by searching Medline, Embase, Emcare, PsycINFO and Cinahl. The databases were searched for terms relating to (rarer dementias) AND (activities of daily living) AND (longitudinal OR cross-sectional studies), using a pre-established protocol registered with the international prospective register of systematic reviews (registration: CRD42021283302).

**Results:** A total of 579 articles were screened for relevant content, of which 20 full-text publications were included in the analysis. Nineteen studies were about rarer dementias on the frontotemporal dementia/primary progressive aphasia spectrum, and one was about posterior cortical atrophy. Long term description of decline was limited to just seven studies following patients for longer than five years. The rate of decline, sequence of symptom onset, and symptom duration were also highlighted.

**Conclusion:** Descriptions of ADL progression were inadequately long term, covering an average of 3.5 years from symptom onset, and lacked phenotypic specificity. The literature disproportionately studied dementias on the frontotemporal dementia spectrum. To facilitate better care, more longitudinal data, quantitative analyses, and development of rarer dementia-specific ADL scales is needed. Given the low prevalence of rarer dementias, big data analyses may never be applicable and so personalised medicine approaches should be pursued, including innovative possibilities in digital biomarkers such as from wearable technology.

## Introduction

Dementia is a clinical syndrome of progressive cognitive decline with impaired social or occupational functioning. There are currently over 850,000 people living with dementia (PLWD) in the UK alone^1^. Of the major neurodegenerative dementias, the most common form is typical Alzheimer’s disease (tAD), which is estimated to account for 50-75% of dementia diagnoses in the UK^1^. Typical Alzheimer’s disease is sporadic, has an older age of onset of symptoms (>65 years) and is characterised by episodic memory loss.

This review considers low prevalence dementias which are estimated to account for less than 10% of total dementia diagnoses in the UK. These ‘rarer dementias’ are often characterised by progressive difficulties with cognitive symptoms other than memory and/or developing at a relatively younger age (pre-65) and/or may be an autosomal dominant disease^2,3^. Figure 1 illustrates the rarer dementias considered in this review and their dominant pathologies.

**Figure 1:**
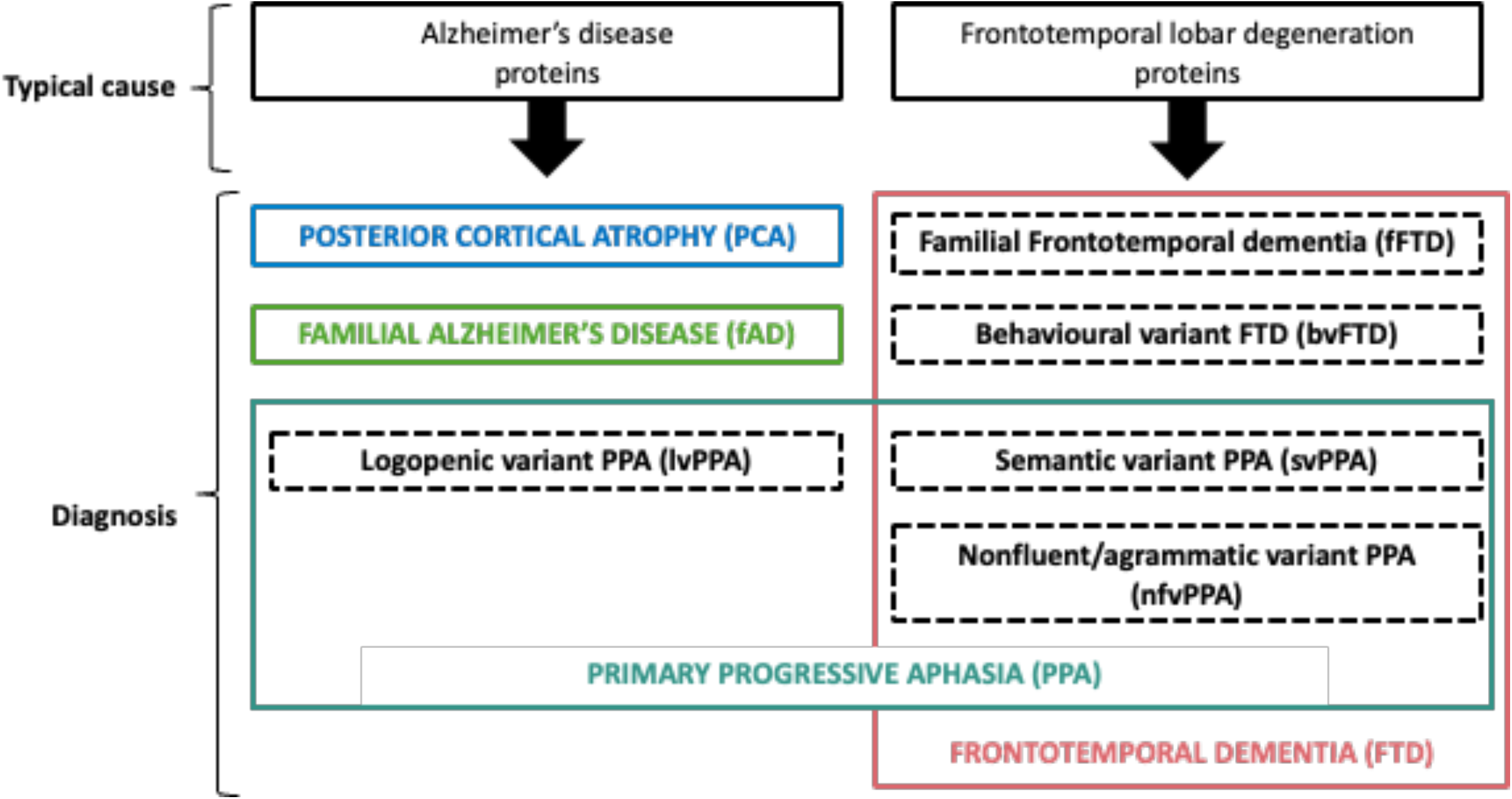
Underlying pathology and subsequent diagnosis for the dementias included in this review. Individuals may have a mixture of multiple dementias.

The term ‘activities of daily living’ (ADL) refers to functional acts that are necessary to live independently, encompassing both ‘basic activities of daily living’ (BADL) such as eating, washing and continence as well as ‘instrumental activities of daily living’ (IADL) such as organising finances, correspondence, and cooking. For tAD, staging scales provide detailed descriptions of how these ADLs change over the course of the disease progression.

For PLWD, care needs can be complex and difficult to address, which reduces their own, and their carers’, health-related quality of life. This is exacerbated by atypical symptoms^4,5^, which are common in the rarer dementias. Many of the rarer dementias are also young onset dementias (YOD). Experiencing dementia at any age can be devastating, but in YOD there are additional concerns and care needs relating to the PLWD’s age, such as work, mortgages, and young children. Furthermore, individuals with YOD often experience a delay to diagnosis which results in difficulties accessing the right care at appropriate times^6^.

Studies on carers’ experiences in the rarer dementias have indicated a need for better education and understanding^7,8^. Understanding of decline in ADLs is important, as this knowledge enables anticipation of care needs, helping carers prepare emotionally and practically for each expected symptom. Objective descriptions of decline are fundamental for evaluating novel care interventions and can be used as outcome measures in clinical trials^9^.

While there have been systematic reviews of activities of daily living in the more prevalent dementias^10^, there is limited systematic synthesis of research into ADLs in people with less common neurodegenerative dementias. This review seeks to understand how ADLs evolve over time, which methods have been used to quantify progression, and what the implications are for care needs in this underserved group of patients.

### Longitudinal ADLs in the most common dementias

Alongside tAD, other prevalent dementias include vascular dementia and Lewy body dementia. Vascular dementia (VaD) refers to conditions of vascular brain damage and is characterised by decline in attention and executive functioning^11^. Lewy body dementia is an umbrella term encompassing dementia with Lewy bodies (DLB) and Parkinson’s disease dementia (PDD): diseases that are pathologically defined by the presence of Lewy bodies, and associated with changes to cognition, parkinsonism, and hallucinations^12,13^. A previous systematic review has addressed ADL decline in the more prevalent neurodegenerative dementias, concentrating on tAD, VaD and PDD^10^. Separate studies have addressed ADLs in DLB, finding that BADLs and IADLs are more impaired early on in DLB compared to tAD^14^, but over time functional changes are comparable^15^.

### Rarer Dementias

Frontotemporal dementia, primary progressive aphasia, posterior cortical atrophy, familial Alzheimer’s disease and familial frontotemporal dementia can be referred to under the collective term of rarer dementia^3^, which can occur concurrently in some people. A summary of the rarer dementias follows.

Frontotemporal Dementia (FTD) is an umbrella term describing clinical syndromes caused by progressive deterioration of networks centred on the frontal and temporal lobes, accounting for 2% of all dementia diagnoses in the UK^1^. The term encompasses behavioural variant FTD (bvFTD)^16^, semantic variant primary progressive aphasia (svPPA) and non-fluent/agrammatic variant primary progressive aphasia (nfvPPA).

Primary progressive aphasia (PPA) is a partially overlapping term with FTD which refers to dementias characterised by initial changes in speech and/or language^17,18^. It’s estimated prevalence is three cases per 100,000^17^ (<1% of all dementia diagnoses in the UK, based on dementia prevalence of 850,000/67.22 million). The three most common variants are svPPA, nfvPPA and logopenic variant PPA (lvPPA)^17,18^.

Posterior cortical atrophy (PCA) is a dementia characterised by changes to visuoperceptual and visuospatial processing^19,20^. The most common pathology associated with PCA is Alzheimer’s disease pathology; however, amongst all individuals with Alzheimer’s disease pathology, fewer than 5% (<2.5-3.75% of all dementia diagnoses in the UK, based on prevalence estimates of tAD) present clinically with PCA^20^.

Familial Alzheimer’s Disease (fAD), and familial FTD (fFTD) are dominantly inherited forms of AD and FTD respectively, with typically younger onset^21,22^. Whilst symptom presentation in these dementias is comparable to the sporadic variants, the fact they are dominantly inherited results in specific care needs relating to the stress of caregivers, who are often the children of patients, having a genetic susceptibility to develop the same condition^23^.

## Methods

### Protocol and registration

The protocol for this systematic review was prepared according to PRISMA guidelines^24^, and registered with PROSPERO (registration: CRD42021283302) prior to data extraction.

### Search strategy and eligibility criteria

A search was conducted across the following five databases: Embase, Medline, Emcare, PsycINFO and Cinahl, from inception up to 7th September 2021. The search terms consisted of three blocks. The first block contained keywords related to longitudinal or cross-sectional studies, the second block contained keywords relating to activities of daily living and the third block contained keywords relating to the rarer dementias. The three blocks were combined with an AND operator.

Studies were included if they (1) were specifically about a dementia whose prevalence was estimated to account for less than 10% of UK dementia. This included: primary progressive aphasia, semantic variant PPA, logopenic variant PPA, non-fluent/agrammatic variant PPA, posterior cortical atrophy, frontotemporal dementia, behavioural variant frontotemporal dementia, familial frontotemporal dementia, familial Alzheimer’s disease. (2) addressed the ADLs of the PLWD. The study had to be primarily about the person living with dementia; in cases where data came from carers the data had to primarily address the needs of the care receiver (i.e. the PLWD). IADLs and BADLs were both considered. (3) were EITHER longitudinal (i.e. reporting on the same individuals over time) OR have a cross-sectional design to examine progression across different groups (i.e. direct comparison of individuals at different disease stages). (4) had a sample size of at least five people living with a rarer dementia, analysed separately to control participants. (5) were published in English in a peer reviewed journal.

Studies were excluded if they were (1) about typical Alzheimer’s Disease, vascular Dementia, Lewy Body Dementia, or non-neurodegenerative Dementias.

### Screening procedure

A three-step procedure was used, first screening studies by title, then by abstract then finally the full text was read and inclusion was determined based on the eligibility criteria. All stages were conducted by the primary reviewer (BT). At all three stages a random 10% of all articles were screened by an independent rater (SM). Disagreements were discussed by the reviewers (BT and SM) and resolved. The studies identified for inclusion were manually citation searched using Google Scholar in March 2022 (by BT and CH) to identify other relevant studies.

### Data extraction

We used a spreadsheet to store extracted data in the following domains: author name(s), publication year, size of sample, study design, length of follow-up (if any), setting (e.g. the clinic that subjects were recruited from), diagnostic criteria, genetic testing, use of in vivo measures to confirm diagnosis (e.g. neuroimaging), confirmation of diagnosis at autopsy, measures of ADLs (including whether the measure was custom developed for the study), method of analysis (e.g. statistical assessment, linear regression), the reported sequence of ADL decline, and the rate of ADL decline.

### Quality rating

A quality assessment of the included literature was undertaken independently by two reviewers (BT and CH) using a quality assessment adapted from the Joanna Briggs Institute critical appraisal tools^25^ with additional questions to address bias specific to the topic^26^, Table 1. Appraisals according to study design are given in the supplementary materials, eTables 1-3.

**Table 1:**
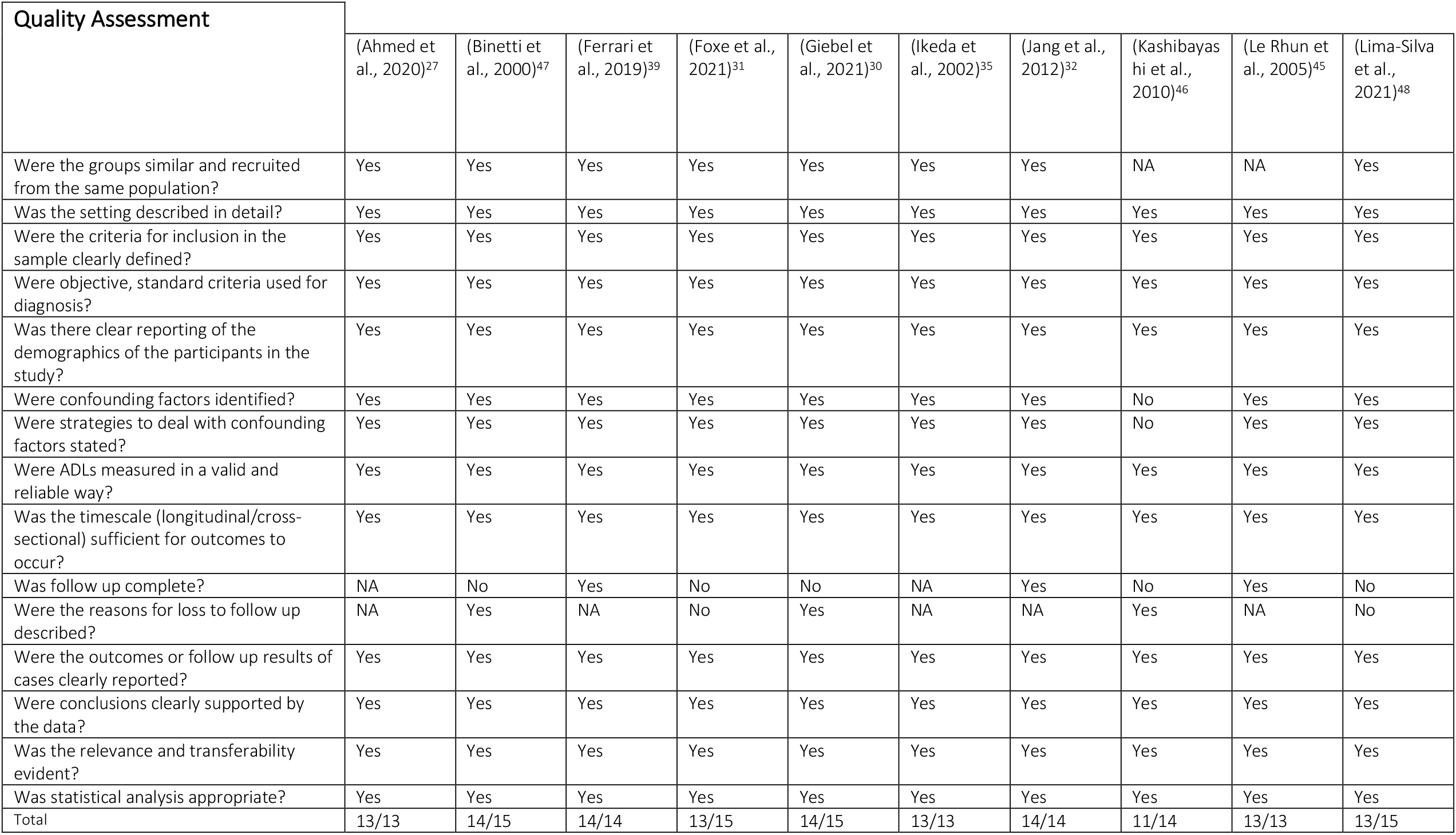
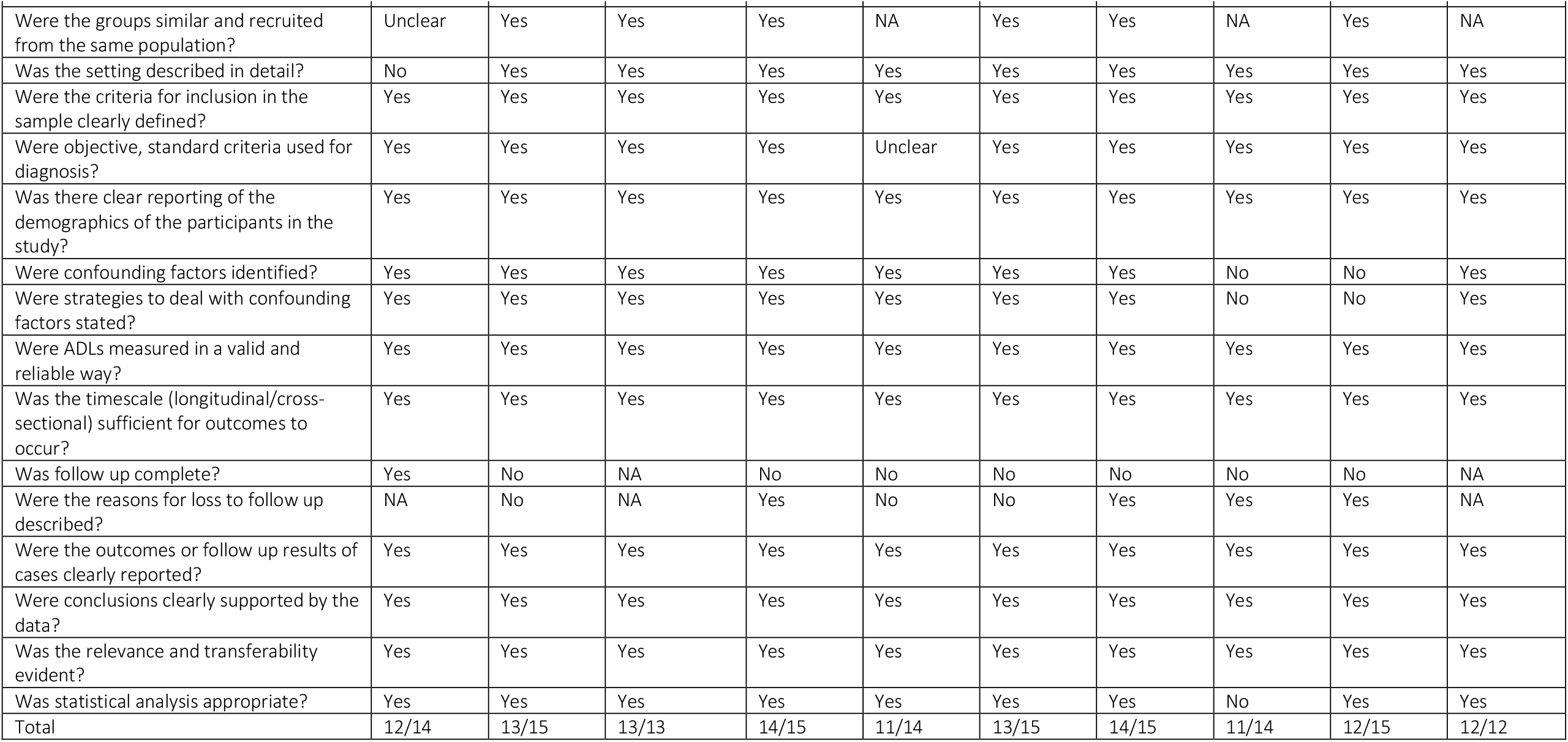
Custom quality assessment of included studies.

## Results

### Selection processes

A total of 864 studies were retrieved, resulting in 579 after removing duplicates. Of these studies 33 were sought for retrieval based on title and abstract, 32 articles were retrievable and read in full by the reviewer with 14 deemed eligible for inclusion. Citation searches of the 14 initial records identified a further six articles. More details can be found in the PRISMA diagram - Figure 2.

**Figure 2:**
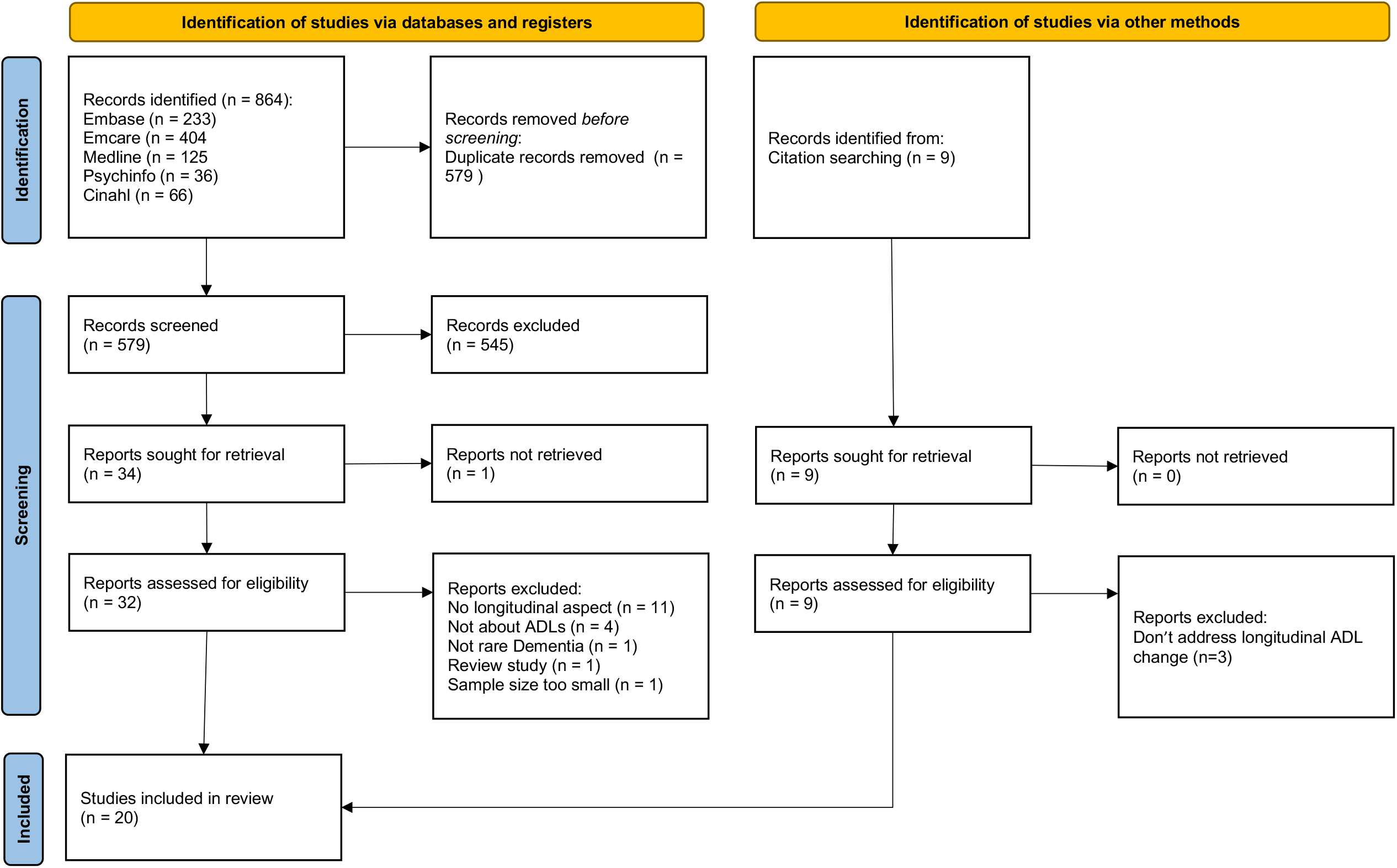
PRISMA flow diagram of the identification of studies.

### Synthesis

We grouped studies according to the syndrome/disease under consideration. To facilitate comparison across dementia variants we use prevailing contemporary diagnostic terminology throughout (for example svPPA rather than semantic dementia — SD). eTable 4 in the supplementary materials contains the original diagnosis of participants in the studies, and the contemporary interpretation.

Table 2 shows the main characteristics and outcomes of interest for each included study. Where given in the original study, p-values were included in the table. Due to the small amount of literature the findings of all studies were reported, with results from higher-quality studies presented first — according to the quality rating in Table 1.

**Table 2:**
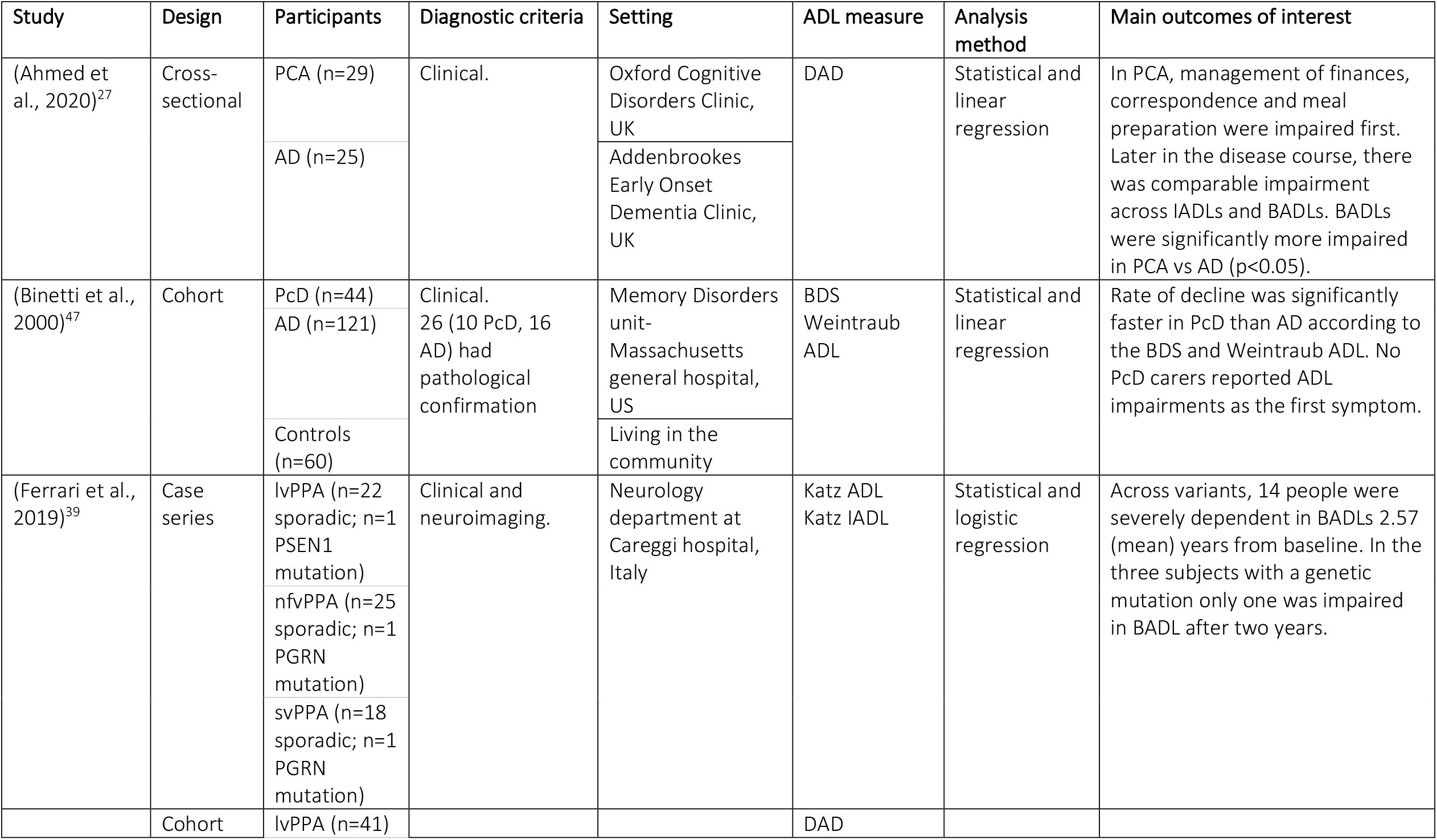
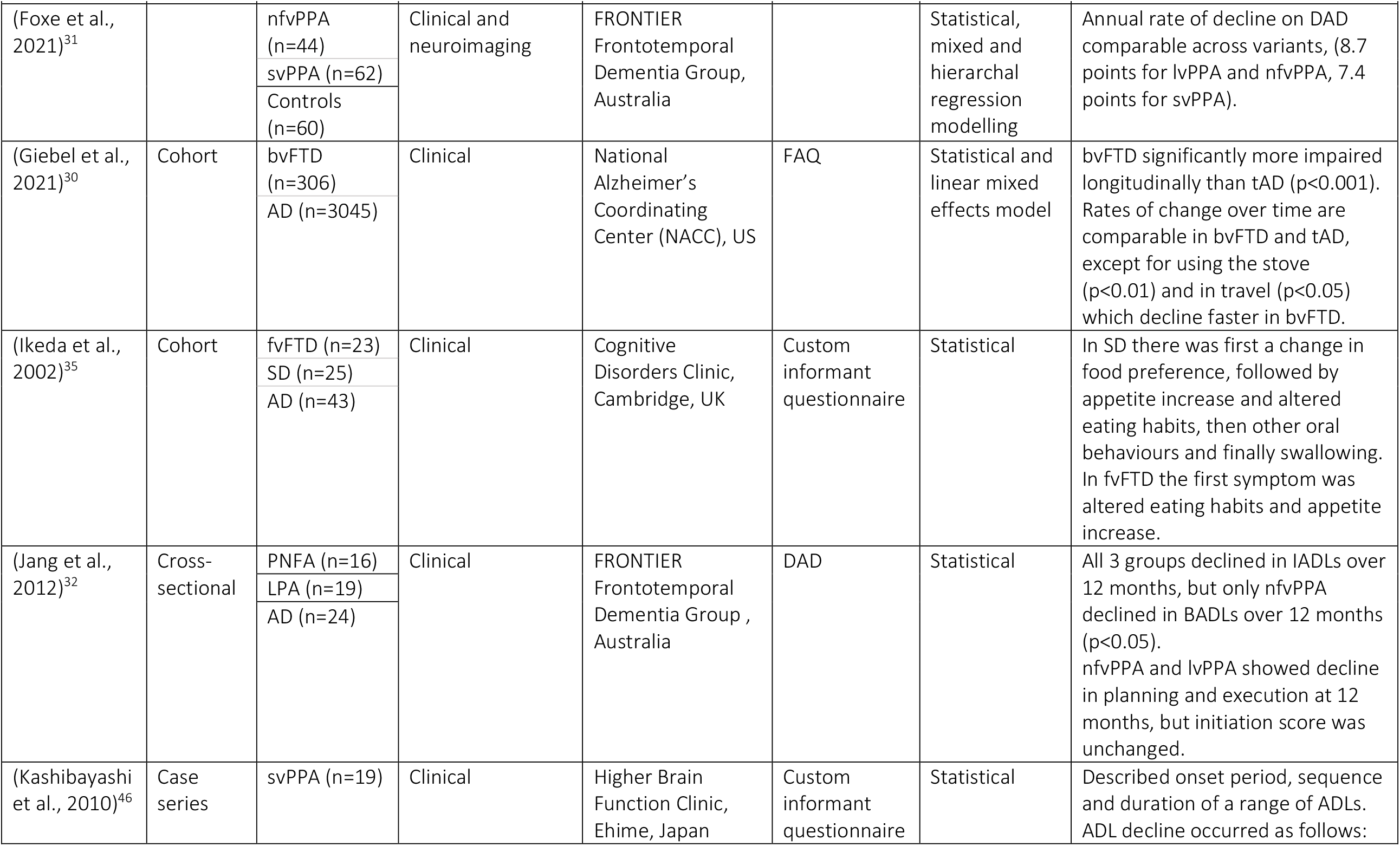
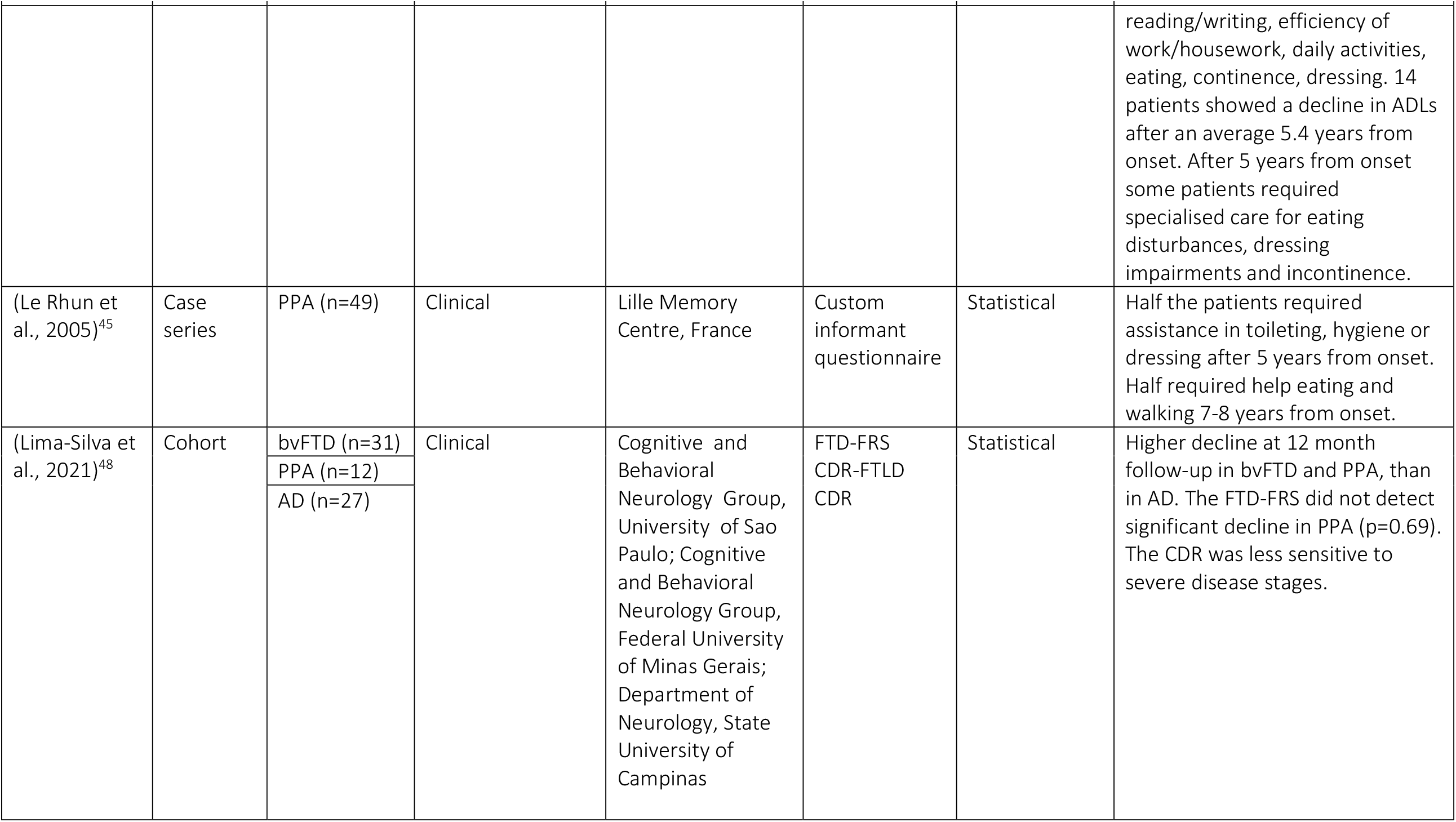
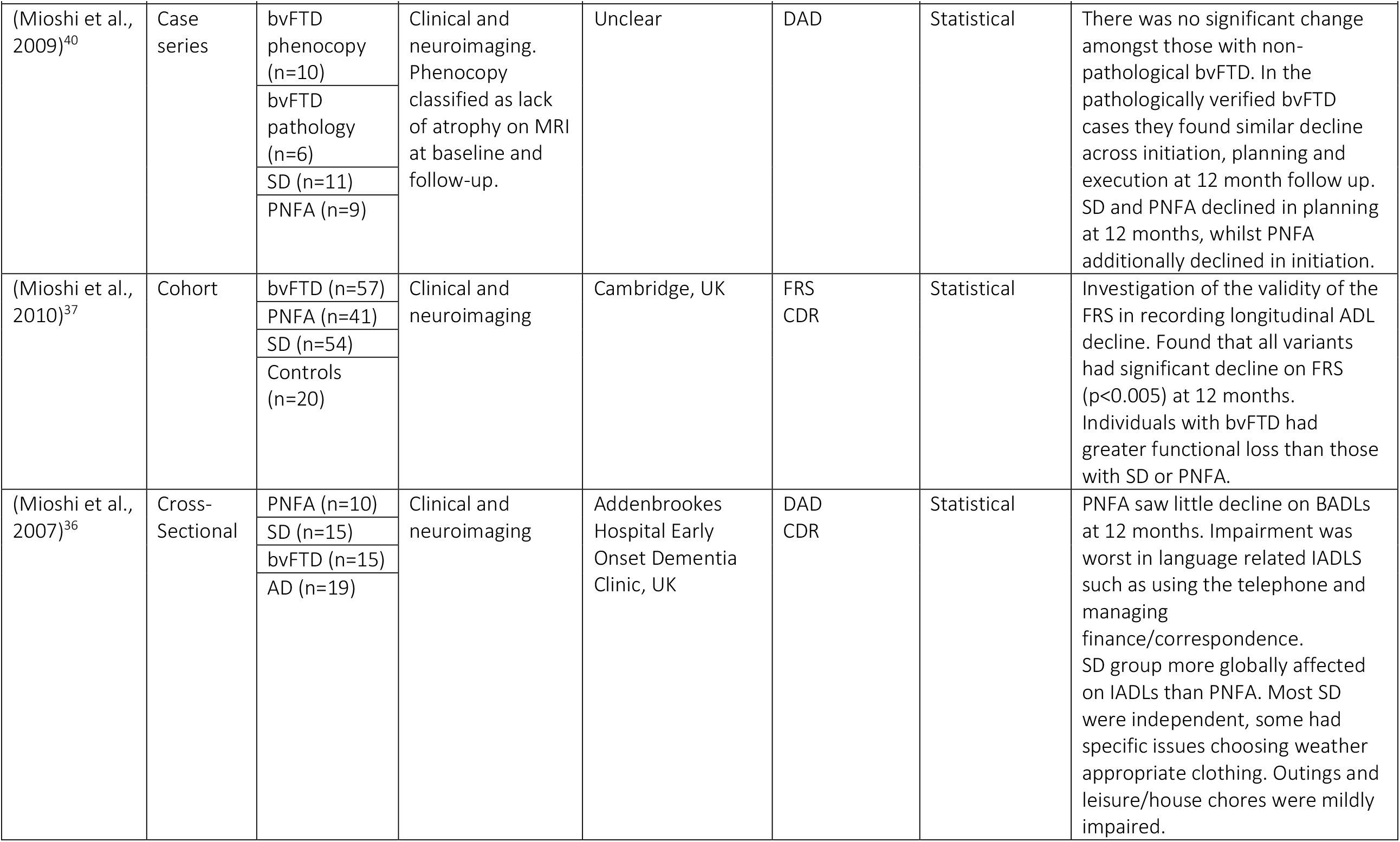
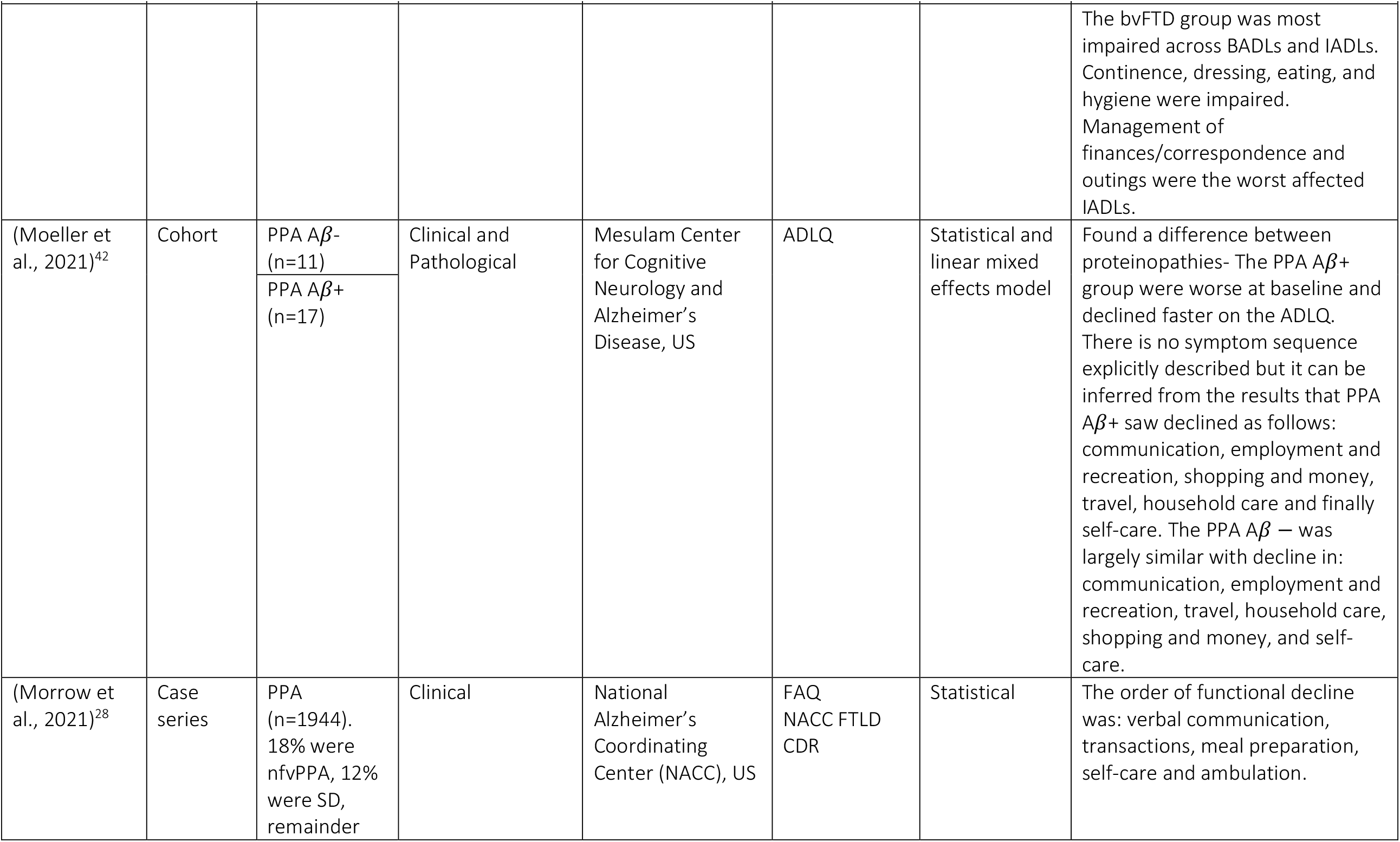
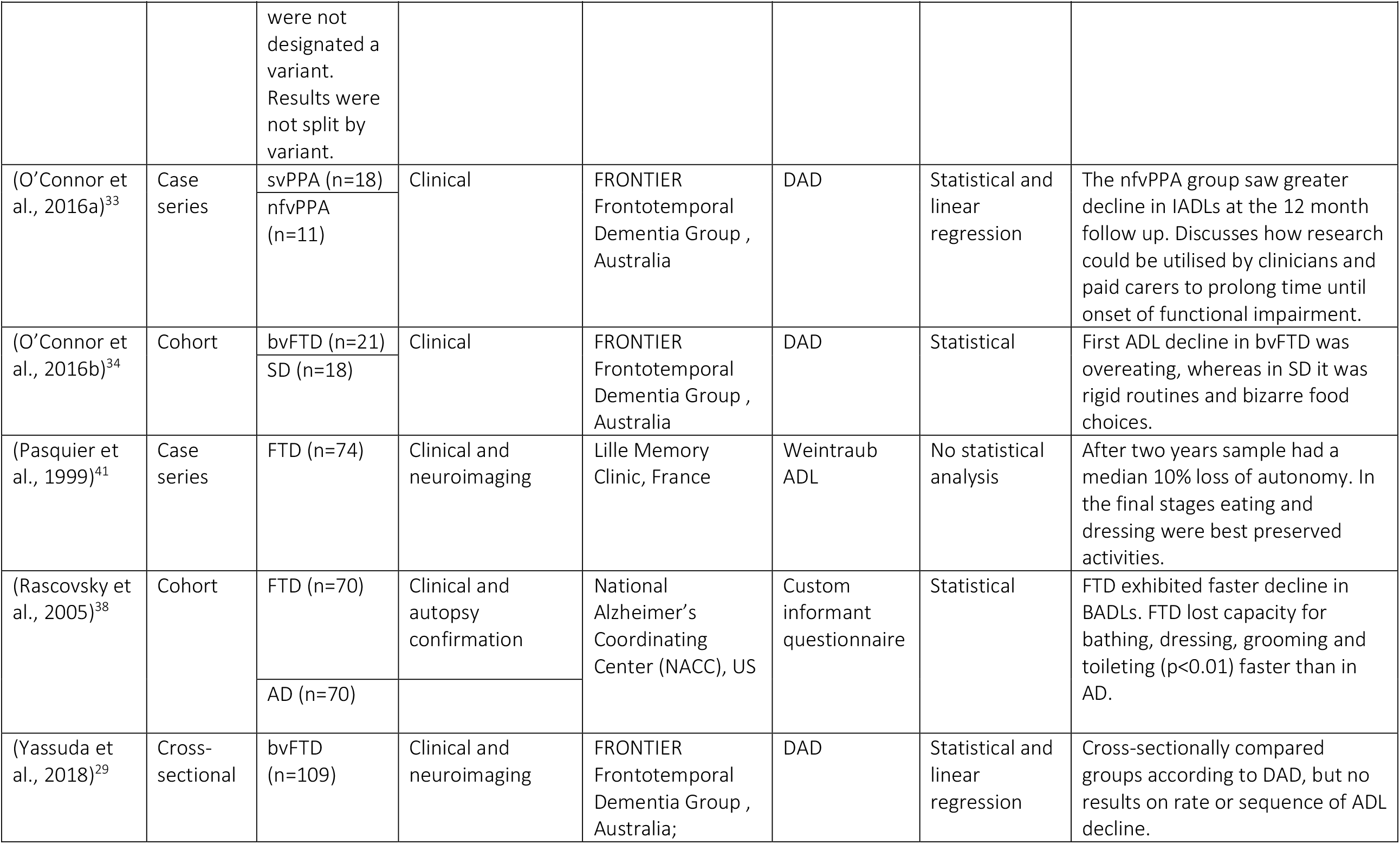
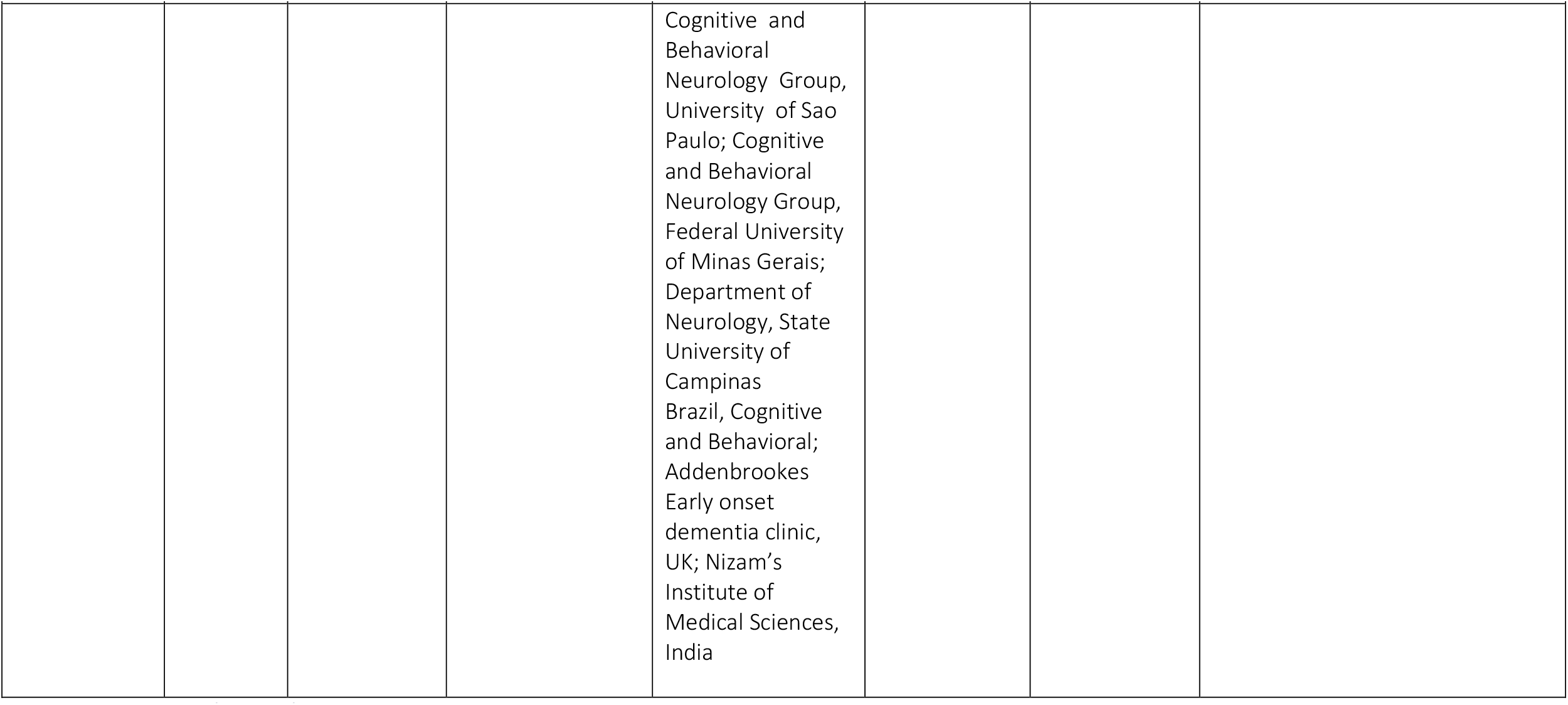
Main outcomes of interest for the 20 studies included in the review. PCA, Posterior Cortical Atrophy; AD, Alzheimer’s Disease; PcD, Pick’s Disease; PPA, Primary Progressive Aphasia; PPA Aβ-, Amyloid beta negative PPA; PPA Aβ+, Amyloid beta positive PPA; lvPPA, logopenic variant PPA; nfvPPA, non-fluent/agrammatic variant PPA; PNFA, progressive non-fluent aphasia; svPPA, semantic variant PPA; SD, semantic dementia; FTD, Frontotemporal Dementia; bvFTD, behavioural variant FTD; fvFTD, frontal variant FTD, PSEN1, presenilin-1; PGRN, progranulin. DAD, Disability Assessment for Dementia; BDS, Blessed Dementia Scale; Weintraub ADL, Weintraub Activities of Daily Living Scale; Katz ADL, The Katz Index of Independence in Activities of Daily Living; IADL, Instrumental Activities of Daily Living Scale; FAQ, Functional Activities Questionnaire.

### Study characteristics and participants

Across the 20 studies there was a total of n=3,309 participants diagnosed with a rarer dementia, some of whom may have been included in multiple studies. Of these participants, the average age at the start of the study was 67.26 years. Study countries included Australia (k=4), Brazil (k=2), France (k=2), Italy (k=1), Japan (k=1), UK (k=4) and the US (k=5). For one study the country was unclear.

Four studies were cross-sectional. Sixteen studies were longitudinal, of which 10 were case series studies and six were cohort studies. In the longitudinal studies, study duration ranged between one and eleven years, with a mean of 3.5 years. Table 3 gives a breakdown of study timeframe versus dementia variant.

**Table 3:**
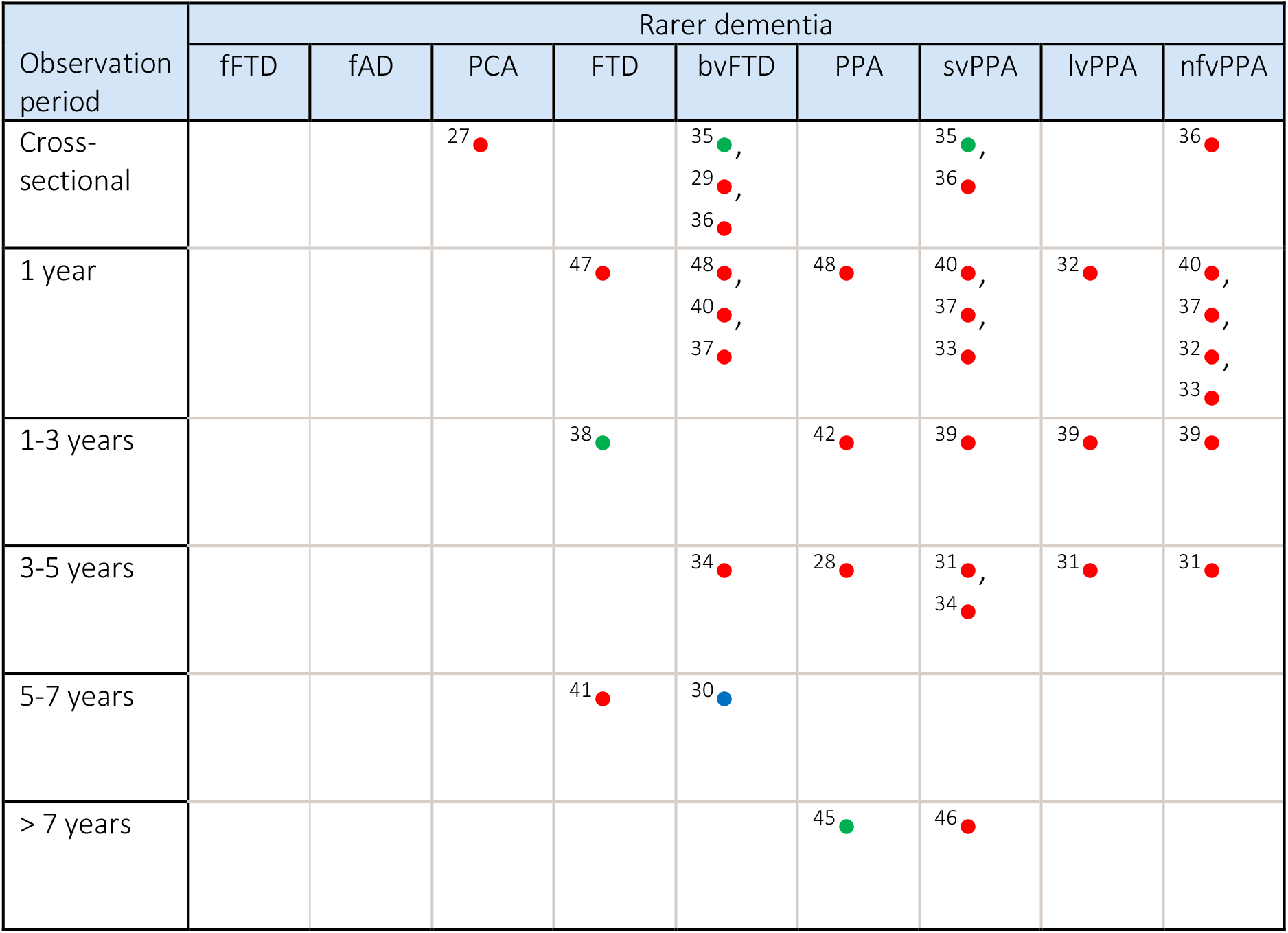
Distribution of studies by PLWD diagnosis versus timeframe of study. Colours indicate what domain of ADL was considered: BADL only, blue; IADL only, green; BADLs and IADLs, red.

Despite the literature search encompassing a broad set of rarer dementia terminology, 19 of the 20 studies focussed on PLWD with a diagnosis in the FTD/PPA spectrum. The remaining study focussed on PCA^27^. Ten studies compared multiple rarer dementias within the FTD/PPA spectrum. As expected for low prevalence diseases small sample sizes were observed, with just three studies exceeding 100 individuals in any single dementia variant^28–30^.

All cohorts studied were recruited through specialist cognitive disorders clinical services. Five studies were conducted using data collected from FRONTIER Frontotemporal Dementia Group, Australia^29,31–34^; four were based in Addenbrookes Hospital, UK^27,35–37^; three used data collected by the National Alzheimer’s Coordinating Center (NACC), US^28,30,38^.

All studies used clinical diagnostic criteria. Some used neuroimaging techniques to confirm diagnosis, such as magnetic resonance imaging^31,36,37,39–42^ or A*β* positron emission tomography^39,42^. One study confirmed pathology at autopsy^38^.

Eight studies used the Disability Assessment for Dementia (DAD)^43^ to measure ADL decline. The DAD consists of 40 items, 17 addressing BADLs and 23 addressing IADLs, each of which are classified as relating to initiation, planning, or organisation. The DAD was primarily developed for assessment of functional decline in tAD^44^. Some studies used custom informant questionnaires, which were tailored to specific dementia variants. One study developed and validated a new FTD rating scale (FTD-FRS)^37^.

All but one study^41^ used statistical methods to describe longitudinal change. Significance tests were used to verify decline; regression was used to predict ADL decline^30,31^. Some studies adjusted for covariates to statistically account for confounding factors^29,30,42,45^.

There was considerable variation in description and assessment of ADL decline. Six studies only collected data at two time points, resulting in symptoms categorised as occurring by baseline, or after 12 months. The most detailed study sequentially ordered eight ADLs, based on data collected over 10 years^46^. Some studies found that the rate of decline was faster in rarer dementias compared to tAD^32,40,47^. Two studies explicitly discussed the implications of the results for addressing care needs^27,37^. Table 3 presents a summary of each study’s relevant findings.

### Results for specific diagnostic variants

#### bvFTD

Eight studies included people living with bvFTD^29,30,34–37,40,48^, including one study of individuals living with frontal variant FTD (fvFTD)^35^. Amongst the five longitudinal studies study duration was in the range of 1-6 years, with a mean of 2.4 years. Two studies identified increased appetite/overeating as an initial symptom in bvFTD^34,35^. The sequence of ADL decline was found to be IADLs followed by BADLs^40^, specifically: initiation > planning > BADLs^36^. Ability to use the stove and to travel were found to decline significantly faster in bvFTD than in tAD, however overall rate of functional decline was comparable^30^. The rate of functional decline was found to be faster in bvFTD compared to svPPA or nfvPPA using a single follow-up twelve months after baseline^37^.

#### svPPA

Nine studies included people living with svPPA^31,33–37,39,40,46^. Amongst the seven longitudinal studies study duration was in the range of 1-10 years, with a mean of 3 years. One study gave a particularly clear description of the sequence of ADL decline, reporting that symptoms occurred as: reading/writing > work/housework > daily activities > eating > continence > dressing^46^. Another study found the first symptom of functional decline was rigid routines and unusual food choices^34^. Annual rate of decline in ADLs was found to be highest in planning, using a single follow-up twelve months after baseline^40^.

There was conflicting evidence as to whether the rate of IADL decline was more pronounced in nfvPPA^33^ or svPPA ^36^. The rate of functional decline was comparable between svPPA and tAD^40^.

#### nfvPPA

Seven studies included people living with nfvPPA^31–33,36,37,39,40^. Amongst the six longitudinal studies study duration was in the range of 1-3.5 years, with a mean of 1.7 years. The initial symptoms of functional decline were found to be language and household chores^32^. There was variation as to whether decline occurred concurrently in BADLs and IADLs using a single follow-up twelve months after baseline^32,40^, or whether decline in BADLs preceded decline in IADLs^36^.

#### lvPPA

Three studies included people living with lvPPA^31,32,39^. All studies were longitudinal, study duration was in the range of 1-3.5 years, with a mean of 2.3 years. This was the least represented of the three most common PPA variants, which might be expected since it is the most recently described ^49^. Decline in IADLs was more significant that decline in BADLs, with marked decline in planning and execution 12 months after baseline ^32^.

#### PCA

One study included people living with PCA^27^. This study was cross-sectional. The sequence of ADL decline was found to be impairment managing finances/correspondence and meal preparation, followed by impairment across BADLs.

### Results for broader diagnostic groups

#### FTD

Three studies included people living with FTD without differentiating between variant^38,41,47^. All studies were longitudinal, study duration was in the range of 1-6 years, with a mean of 2.7 years. Dressing and eating were found to be the best-preserved ADLs ^41^. Annual rates of decline in ADLs found that bathing, grooming, and toileting declined significantly faster in FTD than tAD ^38^.

One study considered Pick’s disease (an older diagnostic term which is broadly synonymous with FTD) with n=10 individuals having pathological confirmation, and n=34 showing “prominent language impairments, and subtle personality changes”^47^. ADL decline was reported to be significantly faster in Pick’s disease compared to tAD^47^.

#### PPA

Four studies included people living with PPA without differentiating by variant^28,42,45,48^. All studies were longitudinal, study duration was in the range of 1-11 years, with a mean of 4.75 years. One study found decline in toileting, hygiene and dressing occurred five years after onset, whilst eating and walking declined seven years after onset^45^. Another study reported the sequence of ADL decline as: verbal communication > transactions > meal preparation > self-care > ambulation^28^.

When the cohort was differentiated according to amyloid-beta status (from florbetapir PET scans), the reported sequence of ADL decline amongst individuals who were amyloid beta positive (A*β*+) was: communication > employment and recreation > shopping and money > travel > household care > self-care^42^. Amongst individuals who were Amyloid beta negative (A*β*-) ‘shopping and money’ declined later.

## Discussion

We provide a systematic review of longitudinal change in ADLs in people diagnosed with a rarer dementia, including studies of the FTD/PPA spectrum but excluding the more common dementias of VaD and LBD which have been reviewed previously^10^. As might be expected, the sequences and rates of ADL decline in rarer dementias differed from observations in tAD, and differed among the rarer dementias. Reported patterns of ADL decline varied between studies within a single dementia syndrome, reflecting either study-level variation in diagnostic specificity, or genuine within-syndrome heterogeneity not captured by single studies. This suggests the need for further studies, ideally with more detailed and dementia-specific descriptions of functional decline, to better understand this intradiagnostic heterogeneity. Some commonalities between studies were observed, with most studies finding that decline in IADLs precedes decline in BADLs, and a subset of symptoms were identified which occurred early in the disease course.

These findings are different to those in the more prevalent dementias. A previous systematic review concerning tAD, VaD, and PDD found that the most affected ADLs are eating and dressing^10^. The review suggests this finding may be influenced by more studies focusing on the latter stages of the disease. Studies on DLB have found greater impairment in BADLs and IADLs initially compared to tAD^14^, but long-term functional changes are comparable^15^.

Of the studies included in this review there was only one outside of the FTD/PPA spectrum. This might be expected from prevalence estimates as FTD is one of the more prevalent rarer dementias, it’s estimated to account for 2% of dementia diagnoses in the UK^1^. Additionally, FTD/PPA findings were dominated by datasets collected from a single dementia research clinic, the FRONTIER Frontotemporal Dementia Group, Australia^29,31–34^.

Just two of the ADL scales used in the studies were specifically developed for the dementia variants they were being used to measure. The use of non-rarer dementia specific staging scales could lead to under reporting of activities of daily living that are variant specific. Since tAD is older age of onset, staging scales developed for tAD fail to measure ADLs more relevant to younger people, meaning that age specific needs are overlooked. The only rarer dementia specific staging scales of ADL decline in the literature were the FTD-FRS and CDR-FTLD, both of which assess FTD decline. The FTD-FRS covers domains of behaviour, outings and shopping, household chores and telephone, finances, medications, meal preparation and eating, and self-care and mobility — a comprehensive assessment of ADL. However, none of the items included specifically pertain to language, potentially limiting its utility for tracking symptom severity in the primary progressive aphasias.

There was variation across studies of the reported sequence in which ADLs declined, as well as variation in the rate of decline, which may be accounted for by diagnostic criteria. Each rarer dementia is itself an umbrella term which conceals heterogeneity in both pathology and disease presentation. The oldest study in this review was published in 1999^41^ — disease definitions and diagnostic criteria have evolved since then. There was a trend for recent studies to classify individuals’ diagnosis more specifically, incorporating neuroimaging results and genetics, thus reducing the likelihood of misdiagnosis.

Longitudinal study duration varied between one and ten years, likely accounting for the variation in reported detail of specific symptoms of ADL decline. Long term studies which follow individuals from diagnosis to death allow for detailed descriptions on when each symptom occurs. Comparatively, studies which collect data at just 12 month follow up from baseline only provide enough information to differentiate between symptoms which occur during that year, and those that occur later. Due to the short timescale of studies, the existing literature does not appropriately nor equally represent activities of daily living in all (early, middle, and late) stages of decline.

There was very limited practical discussion of how findings could be used to support care, perhaps due in part to the insufficient coverage of all stages of decline described above. Staging scales that provide detailed descriptions of functional decline can enable carers to make legal, financial, care planning and housing decisions, thus reducing carer burden^7,8^, and supporting independence for PLWD. Furthermore, findings could inform support groups which create networks of individuals with similar rarer diagnoses, facilitating both formal teaching, and informal peer-to-peer education.

## Limitations

There are limitations to this review. In excluding studies that did not explicitly address ADL decline as a primary focus, we may have overlooked studies where ADL decline was secondary to a description of progression in terms of neuroimaging or psychosocial symptoms. This review covered a range of different dementias which limited the cross comparison of studies. Four studies were cross-sectional, and amongst the longitudinal studies follow-up time varied, precluding a meta-analysis.

### Recommendations and Future Directions

To better understand ADL decline in the dementias, especially the rarer dementias, requires more longitudinal data, quantitative analyses, and development of dementia-specific ADL staging scales. Future work could employ emerging research into wearables^50^ to collect longitudinal data over a wider range of ADLs specifically selected for their relevance to dementia variant, at more frequent and regular intervals. Wearables would also facilitate verification of self-reported data, which may overestimate or underestimate abilities.

Future work must address ADL decline outside of the FTD/PPA spectrum. There were no studies on autosomal-dominantly inherited dementias. fAD and fFTD are typically young onset dementias^21,22^; as such they have additional age-related needs, including complications around employment and parenting^8^. Hence, it would be interesting to see a description of ADL decline specific to these genetic/familial forms of rarer dementias.

Furthermore, future studies need to provide more detailed and longitudinal descriptions of ADL decline across all the specific rarer dementia variants. Understanding functional decline in rarer dementias is integral for developing personalised care. The implications for how these findings could help in care could be more explicitly addressed, for example by discussing how results could be translated into useful clinical staging scales.

Staging scales for tAD, like those used in the literature to measure decline, fail to include all the symptoms relevant to each rarer form of dementia. The development of staging scales specific to rarer dementias could better account for heterogeneity and would be a useful tool in providing additional information about the expected disease course for carers and PLWD. This work could utilise computational methods, which are increasingly being used to quantify progression in dementia. The small sample sizes in rarer dementia research limits the application of machine learning, however slightly larger datasets could be combined with computational modelling techniques to gain further insight into ADL decline.

## Conclusions

Our review demonstrates the need for more comprehensive and specific descriptions of longitudinal decline in activities of daily living in rarer dementias, especially outside the FTD/PPA spectrum. The lack of knowledge necessitates studies with more longitudinal follow up, and the development and application of emerging computational techniques to rigorously quantify change. There should be a clear direction on how these descriptions can be used to inform care needs, for example in developing variant specific staging scales.

## Data Availability

Systematic review - not applicable.

