## Supplemental materials for "How do the activities of daily living decline in people living with rarer dementias? A systematic review"

### Supplementary materials

#### Search strategy

A search was carried out in Medline, Embase, Emcare, PsychINFO and Cinahl in September 2021. The searches are given below.

Medline

1. Epidemiologic studies/ OR exp Case control studies/ OR exp Cohort studies/ OR case control.tw. OR (cohort adj (study OR studies)).tw. OR cohort analy$.tw. OR (follow up adj (study OR studies)).tw. OR (observational adj (study OR studies)).tw. OR longitudinal.tw. OR retrospective.tw. OR cross sectional.tw. OR cross-sectional studies/
2. primary progressive aphasia$.tw. OR exp Aphasia, primary progressive/ OR (semantic dementia OR svPPA).tw. OR (logopenic aphasia OR lvPPA).tw. OR (progressive nonfluent aphasia OR nfvPPA).tw. OR (posterior cortical atrophy OR bensons syndrome).tw. OR frontotemporal dementia.tw. OR exp Frontotemporal Lobar Degeneration/ OR (frontotemporal lobar degeneration OR frontotemporal degeneration OR Picks disease).tw. OR fFTD.tw. OR familial frontotemporal dementia.tw. OR (behavioral variant frontotemporal dementia OR behavioural variant frontotemporal dementia).tw. OR familial alzheimers disease.tw. OR atypical alzheimers disease.tw. OR frontal variant alzheimers disease.tw. OR rare dementia$.tw.
3. (carer$ OR "care need$" OR "care partner$" OR caregiver$).mp. OR Caregivers/ OR "Activities of Daily Living"/ OR activities of daily living.tw. OR (IADLs OR IADL).tw. OR (ADLs OR ADL).tw.
4. 1 AND 2 AND 3 limited to English

Embase

1. Clinical study/ OR exp case control study/ OR Family study/ OR Longitudinal study/ OR Retrospective study/ OR (Prospective study/ NOT Randomized controlled trials/) OR Cohort analysis/ OR (Cohort adj (study OR studies)).mp. OR (Case control adj (study OR studies)).tw. OR (follow up adj (study OR studies)).tw. OR (observational adj (study OR studies)).tw. OR (epidemiologic$ adj (study OR studies)).tw. OR (cross sectional adj (study OR studies)).tw.
2. primary progressive aphasia$.tw. OR exp primary progressive aphasia/ OR (semantic dementia OR svPPA).tw. OR (logopenic aphasia OR lvPPA).tw. OR (progressive nonfluent aphasia OR nfvPPA).tw. OR (posterior cortical atrophy OR bensons syndrome).tw. OR frontotemporal dementia.tw. OR exp frontotemporal dementia/ OR (frontotemporal degeneration OR frontotemporal lobar degeneration OR Picks disease).tw. OR (behavioural variant frontotemporal dementia OR behavioral variant frontotemporal dementia).tw. OR (familial frontotemporal dementia OR fFTD).tw. OR familial alzheimers disease.tw. OR atypical alzheimers disease.tw. OR frontal variant alzheimers disease.tw. OR rare dementia$.tw.
3. (carer$ OR "care needs" OR "care partner$" OR caregiver$).mp. OR Caregivers/ OR daily life activity/ OR activities of daily living.tw. OR (IADLs OR IADL).tw. OR (ADLs OR ADL).tw.

4. 1 AND 2 AND 3 limited to English

Emcare

1. exp Clinical study/ OR exp case control study/ OR Family study/ OR Longitudinal study/ OR Retrospective study/ OR (Prospective study/ NOT Randomized controlled trials/) OR
   Cohort analysis/ OR (Cohort adj (study OR studies)).mp. OR (Case control adj (study OR studies)).tw. OR (follow up adj (study OR studies)).tw. OR (observational adj (study OR studies)).tw. OR (epidemiologic$ adj (study OR studies)).tw. OR (cross sectional adj (study OR studies)).tw.
2. primary progressive aphasia$.tw. OR exp primary progressive aphasia/ OR (semantic dementia OR svPPA).tw. OR (logopenic aphasia OR lvPPA).tw. OR (progressive nonfluent aphasia OR nfvPPA).tw. OR (posterior cortical atrophy OR bensons syndrome).tw. OR exp brain cortex atrophy/ OR frontotemporal dementia.tw. OR exp frontotemporal dementia/ OR (frontotemporal degeneration OR frontotemporal lobar degeneration OR Picks disease).tw. OR (behavioural variant frontotemporal dementia OR behavioral variant frontotemporal dementia).tw. OR (familial frontotemporal dementia OR fFTD).tw.
   familial alzheimers disease.tw. OR atypical alzheimers disease.tw. OR frontal variant alzheimers disease.tw. OR rare dementia$.tw.
3. (carer$ OR "care needs" OR "care partner$" OR caregiver$).mp. OR Caregivers/
   daily life activity/ OR activities of daily living.tw. OR (ADL OR ADLs).tw. OR (IADLs OR IADL).tw.
4. 1 AND 2 AND 3 limited to English

PsycINFO

1. Cohort analysis/ OR (Cohort adj (study OR studies)).mp. OR (Case control adj (study OR studies)).tw. OR (follow up adj (study OR studies)).tw. OR (observational adj (study OR studies)).tw. OR (epidemiologic$ adj (study OR studies)).tw. OR (cross sectional adj (study OR studies)).tw. OR longitudinal.tw. OR exp retrospective studies/
   exp longitudinal studies/ OR followup studies/ OR
2. primary progressive aphasia$.tw. OR (semantic dementia OR svPPA).tw. OR (logopenic aphasia OR lvPPA).tw. OR (progressive nonfluent aphasia OR nfvPPA).tw. OR exp semantic dementia/ OR (posterior cortical atrophy OR bensons syndrome).tw. OR frontotemporal dementia.tw. OR (frontotemporal lobar degeneration OR frontotemporal degeneration OR Picks disease).tw. OR fFTD.tw. OR familial frontotemporal dementia.tw. OR (behavioral variant frontotemporal dementia OR behavioural variant frontotemporal dementia).tw. OR familial alzheimers disease.tw. OR atypical alzheimers disease.tw. OR frontal variant alzheimers disease.tw. OR rare dementia$.tw.
3. (carer$ OR "care need$" OR "care partner$" OR caregiver$).mp. OR caregivers/ OR exp "Activities of Daily Living"/ OR activities of daily living.tw. OR (IADLs OR IADL).tw. OR (ADLs OR ADL).tw.
4. 1 AND 2 AND 3 limited to English

Cinahl

1. IADLs OR IADL OR ADLs OR ADL OR "activities of daily living" OR (MH "Altered Activities of Daily Living (NANDA)") OR (MH "Self Care: Activities of Daily Living (Iowa NOC)") OR (MH "Self-Care: Instrumental Activities of Daily Living (Iowa NOC)") OR (MH "Activities of Daily Living (Saba CCC)") OR (MH "Instrumental Activities of Daily Living Alteration (Saba CCC)") OR (MH "Instrumental Activities of Daily Living (Saba CCC)") OR (MH "Activities of Daily Living (Saba CCC)") OR (MH "Activities of Daily Living") OR (MH "Caregivers") OR carer* OR "care need*" OR "care partner*" OR "caregiver*"
2. "rare dementia*" OR "frontal variant alzheimers disease" OR "atypical alzheimers disease" OR "behavioural variant frontotemporal dementia" OR "behavioral variant frontotemporal dementia" OR "frontotemporal lobar degeneration" OR "frontotemporal degeneration" OR "picks disease" OR "primary progressive aphasia" OR nfvPPA OR "semantic dementia" OR svPPA OR "logopenic dementia" OR lvPPA OR "familial alzheimer's disease" OR "familial frontotemporal dementia" OR fFTD OR (MH "Frontotemporal Dementia+") OR "frontotemporal dementia" OR "posterior cortical atrophy" OR "bensons syndrome" OR "primary progressive aphasia"
3. "observational study" OR "observational studies" OR "cohort study" OR "cohort studies" OR (MH "Cross sectional studies") OR (MH "Nonconcurrent prospective studies") OR (MH "Correlational studies") OR (MH "Case Control Studies+") OR (MH "Prospective studies")
4. 1 AND 2 AND 3 limited to English

### Study selection

In total 32 papers reached the final stage of the filtering process, which were then included or excluded based on whether they met every point of the criteria. For example *Diehl-Schmid J, Richard-Devantoy S, Grimmer T, Förstl H, Jox R. Behavioral variant frontotemporal dementia: advanced disease stages and death. A step to palliative care. International journal of geriatric psychiatry. 2017 Aug;32(8):876-81.* was excluded at this point, as whilst it addressed ADLs in a rarer dementia, it was not longitudinal. Other studies failed to meet the criteria as they focussed on psychosocial domains rather than ADLs, for example *Bak TH, Crawford LM, Berrios G, Hodges JR. Behavioural symptoms in progressive supranuclear palsy and frontotemporal dementia. Journal of Neurology, Neurosurgery & Psychiatry. 2010 Sep 1;81(9):1057-9.*

### Tabulating and visually displaying study results

Tables and figures were made in Word using the extracted data recorded in the spreadsheet.

| COHORT | Author (year) | | | | | |
|---|---|---|---|---|---|---|
| | (Binetti et al., 2000)[47] | (Foxe et al., 2021)[31] | (Giebel et al., 2021)[30] | (Jang et al., 2012)[32] | (Rascovsky et al., 2005)[38] | (Lima-Silva et al., 2021)[48] |
| Were the two groups similar and recruited from the same population? | Yes | Yes | Yes | Yes | Yes | Yes |
| Were the exposures measured similarly to assign people to both exposed and unexposed groups? | Yes | Yes | Yes | Yes | Yes | Yes |
| Was the exposure measured in a valid and reliable way? | Yes | Yes | Yes | Yes | Yes | Yes |

| | | | | | | |
|---|---|---|---|---|---|---|
| Were confounding factors identified? | Yes | Yes | Yes | Yes | No | Yes |
| Were strategies to deal with confounding factors stated? | Yes | Yes | Yes | Yes | No | Yes |
| Were the groups/participants free of the outcome at the start of the study (or at the moment of exposure)? | Yes | Yes | Yes | Yes | Yes | Yes |
| Were the outcomes measured in a valid and reliable way? | Yes | Yes | Yes | Yes | Yes | Yes |
| Was the follow up time reported and sufficient to be long enough for outcomes to occur? | Yes | Yes | Yes | Yes | Yes | Yes |
| Was follow up complete, and if not, were the reasons to loss to follow up described and explored? | No | No | Yes | No | Yes | Yes |
| Were strategies to address incomplete follow up utilized? | No | No | No | No | No | NA |
| Was appropriate statistical analysis used? | Yes | Yes | Yes | Yes | Yes | Yes |
| Total (/11) | 9 | 9 | 10 | 9 | 8 | 10 |

*eTable 1: Joanna Briggs Institute critical assessment of cohort studies.*

| CROSS-SECTIONAL | Author (year) | | | |
|---|---|---|---|---|
| | (Ahmed et al., 2020)[27] | (Ikeda et al., 2002)[35] | (Mioshi et al., 2007)[36] | (Yassuda et al., 2018)[29] |
| Were the criteria for inclusion in the sample clearly defined? | Yes | Yes | Yes | Yes |
| Were the study subjects and the setting described in detail? | Yes | Yes | Yes | Yes |
| Was the exposure measured in a valid and reliable way? | Yes | Yes | Yes | Yes |
| Were objective, standard criteria used for measurement of the condition? | Yes | Yes | Yes | Yes |
| Were confounding factors identified? | Yes | Yes | Yes | Yes |
| Were strategies to deal with confounding factors stated? | Yes | Yes | Yes | Yes |
| Were the outcomes measured in a valid and reliable way? | Yes | Yes | Yes | Yes |
| Was appropriate statistical analysis used? | Yes | Yes | Yes | Yes |
| Total (/8) | 8 | 8 | 8 | 8 |

*eTable 2:Joanna Briggs Institute critical assessment of cross-sectional studies.*

| CASE SERIES | Author (year) | | | | | | | | | |
|---|---|---|---|---|---|---|---|---|---|---|
| | (Ferrari et al., 2019)[39] | (Kashibayashi et al., 2010)[46] | (Le Rhun et al., 2005)[45] | (Mioshi et al., 2009)[40] | (Mioshi et al., 2010)[37] | (Moeller et al., 2021)[42] | (Morrow et al., 2021)[28] | (O'Connor et al., 2016a)[33] | (O'Connor et al., 2016b)[34] | (Pasquier et al., 1999)[41] |
| Were there clear criteria for inclusion in the case series? | Yes | Yes | Yes | Yes | Yes | Yes | Yes | Yes | Yes | Yes |
| Was the condition measured in a standard, reliable way for all participants included in the case series? | Yes | Yes | Yes | Yes | Yes | Yes | Yes | Yes | Yes | Yes |
| Were valid methods used for identification of the condition for all participants included in the case series? | Yes | Yes | Yes | Yes | Yes | Yes | Yes | Yes | Yes | Yes |
| Did the case series have consecutive inclusion of participants? | Unclear | Yes | Yes | No | Yes | Unclear | Unclear | Yes | Unclear | Yes |
| Did the case series have complete inclusion of participants? | No | No | Yes | No | No | Unclear | Unclear | No | No | Yes |
| Was there clear reporting of the demographics of the participants in the study? | Yes | Yes | Yes | Yes | Yes | Yes | Yes | Yes | Yes | Yes |
| Was there clear | Yes | Yes | Yes | Yes | Yes | Yes | Yes | Yes | Yes | Yes |

| | | | | | | | | | | |
|---|---|---|---|---|---|---|---|---|---|---|
| reporting of clinical information of the participants? | | | | | | | | | | |
| Were the outcomes or follow up results of cases clearly reported? | Yes | Yes | Yes | Yes | Yes | Yes | Yes | Yes | Yes | Yes |
| Was there clear reporting of the presenting site(s)/clinic(s) demographic information? | No | No | No | No | Yes | Yes | No | No | Yes | Yes |
| Was statistical analysis appropriate? | Yes | Yes | Yes | Yes | Yes | Yes | Yes | Yes | Yes | Yes |
| Total (/10) | 7 | 8 | 9 | 7 | 9 | 8 | 7 | 8 | 8 | 10 |

*eTable 3: Joanna Briggs Institute critical assessment of case series studies.*

| Study | Diagnosis of Participants in study | Contemporary Diagnosis of Participants in study |
|---|---|---|
| (Ahmed et al., 2020)[27] | PCA | PCA |
| | AD | AD |
| (Binetti et al., 2000)[47] | Pick's Disease (PcD) | FTD |
| | AD | AD |
| (Ferrari et al., 2019)[39] | lvPPA | lvPPA |
| | nfvPPA | nfvPPA |
| | svPPA | svPPA |
| (Foxe et al., 2021)[31] | lvPPA | lvPPA |
| | nfvPPA | nfvPPA |
| | svPPA | svPPA |
| (Giebel et al., 2021)[30] | bvFTD | bvFTD |
| | AD | AD |
| (Ikeda et al., 2002)[35] | Frontal variant FTD (fv-FTD) | bvFTD |
| | Semantic dementia (SD) | svPPA |
| | AD | AD |
| (Jang et al., 2012)[31] | Progressive non-fluent aphasia (PNFA) | nfvPPA |

| Study | Diagnosis of Participants in study | Contemporary Diagnosis of Participants in study |
|---|---|---|
| | Logopenic progressive aphasia (LPA) | lvPPA |
| | AD | AD |
| (Kashibayashi et al., 2010)[45] | Semantic dementia (SD) | svPPA |
| (Le Rhun et al., 2005)[44] | PPA | PPA |
| (Lima-Silva et al., 2021)[47] | bvFTD | bvFTD |
| | PPA | PPA |
| | AD | AD |
| (Mioshi et al., 2009)[39] | bvFTD | bvFTD |
| | Semantic dementia (SemDem) | svPPA |
| | Progressive non-fluent aphasia (PNFA) | nfvPPA |
| (Mioshi et al., 2010)[36] | bvFTD | bvFTD |
| | Progressive non-fluent aphasia (PNFA) | nfvPPA |
| | Semantic dementia (SemD) | svPPA |
| (Mioshi et al., 2007)[35] | Progressive non-fluent aphasia (PNFA) | nfvPPA |
| | Semantic dementia | svPPA |
| | bvFTD | bvFTD |
| | AD | AD |
| (Moeller et al., 2021)[41] | PPA | PPA |
| (Morrow et al., 2021)[27] | PPA | PPA |
| (O'Connor et al., 2016a)[32] | svPPA | svPPA |
| | nfvPPA | nfvPPA |
| (O'Connor et al., 2016b)[33] | bvFTD | bvFTD |
| | svPPA | svPPA |
| (Pasquier et al., 1999)[40] | FTD | FTD |
| (Rascovsky et al., 2005)[37] | FTD | FTD |
| | AD | AD |
| (Yassuda et al., 2018)[28] | bvFTD | bvFTD |

*eTable 4: We coded frontal variant FTD (fv-FTD) as bvFTD since the study identified it as the frontotemporal dementia variant with reported behavioural changes[34]. The terminology used reflects changing diagnostic criteria; Pick's disease is now used to refer to the pathological presence of Pick's bodies which can only be confirmed at autopsy.*

*PCA, Posterior Cortical Atrophy; AD, Alzheimer's Disease; PPA, Primary Progressive Aphasia; lvPPA, logopenic variant PPA; nfvPPA, non-fluent/agrammatic variant PPA; PNFA, progressive non-fluent aphasia; svPPA, semantic variant PPA; FTD, Frontotemporal Dementia; bvFTD, behavioural variant FTD; fvFTD, frontal variant FTD.*
